# Agreement between state registry, health record, and self-report of influenza vaccination

**DOI:** 10.1101/2021.01.22.21249594

**Authors:** Joshua G. Petrie, Helene Fligiel, Lois Lamerato, Emily T. Martin, Arnold S. Monto

**Affiliations:** Department of Epidemiology, University of Michigan School of Public Health, Ann Arbor; Department of Public Health Sciences, Henry ford Health System, Detroit

**Keywords:** Influenza, Influenza Vaccination, Vaccine Registry, Self-Report, Vaccine Effectiveness

## Abstract

**Background:** Documentation of influenza vaccination, including the specific product received, is critical to estimate annual vaccine effectiveness (VE).

**Methods:** We assessed performance of the Michigan Care Improvement Registry (MCIR) in defining influenza vaccination status relative to documentation by provider records or self-report among subjects enrolled in a study of influenza VE from 2011 through 2019.

**Results:** The specificity and positive predictive value of MCIR were high; however, >10% of vaccinations were identified only by other sources each season. The proportion of records captured by MCIR increased from a low of 67% in 2013-2014 to a high of 89% in 2018-2019, largely driven by increased capture of vaccination among adults.

**Conclusions:** State vaccine registries, such as MCIR, are important tools for documenting influenza vaccination, including the specific product received. However, incomplete capture suggests that documentation from other sources and self-report should be used in combination with registries to reduce misclassification.

---

Rapid evolution of influenza viruses leads to loss of population immunity resulting in annual epidemics[1]. Because of this influenza virus antigenic drift, the strain composition of influenza vaccines is evaluated twice annually (once for the Northern Hemisphere, and once for the Southern) often resulting in updates. Annual observational studies are carried out in many countries to determine the actual effectiveness of the administered vaccine which can vary widely from year to year, largely related to the match between the vaccine and circulating virus strains[2]. These studies depend on accurate determination of influenza vaccination status for unbiased estimation of influenza vaccine effectiveness. Methods for determining influenza vaccination status vary across studies. Self-report of vaccination has previously been shown to have relatively high sensitivity and specificity relative to comprehensive vaccination registries[3–7]. However, documentation of vaccination is typically required to ascertain details about the specific vaccine product. A variety of influenza vaccine products are now licensed for use in the United States with important differences in the type of vaccine (inactivated vs live-attenuated), number of strains included (trivalent vs quadrivalent), method of production (egg grown, cell-culture grown, recombinant), dose (high dose vs standard dose), and inclusion of an adjuvant[8]. Comparison of the effectiveness of these various products requires complete and accurate documentation of the vaccines that individuals receive. Unfortunately, the performance of sources of vaccination documentation can vary and may not outperform self-report[9].

The immunization registry for the state of Michigan (Michigan Care Improvement Registry [MCIR]) was created in 1998 with mandated reporting of all vaccinations administered in the state to persons younger than 20 years of age[10]. In 2006, the registry was expanded to accommodate vaccination records for people of all ages, although only the reporting of vaccinations for children remained mandatory. We compared the performance of MCIR in defining influenza vaccination status relative to self-report and vaccine provider records.

## METHODS

### Study Population

This study included data collected as part of the US Flu Vaccine Effectiveness Network study at the Michigan participating site during eight consecutive influenza seasons from 2011-2012 through 2018-2019. Participants were identified when they presented for treatment of acute respiratory illness to Michigan Medicine or Henry Ford Health System outpatient clinics throughout Southeast Michigan. Patients ≥6 months of age were eligible for enrollment if they were seeking care for an acute respiratory illness with a cough of 7 or fewer days duration. Participant characteristics, including age, sex, race, and Hispanic ethnicity were determined by self-report during an enrollment interview. Informed consent was obtained from participants over age 18 or the parent/guardian of participants under age 18; participants between ages 7-17 additionally provided verbal assent for participation. The study was reviewed and approved by the IRBs at the University of Michigan Medical School and Henry Ford Health System.

### Vaccination Status

Participants, or their parent/guardian, reported their influenza vaccination status at enrollment. If vaccinated, they were also asked to report the date and location of vaccine receipt. Participants were defined vaccinated by self-report if they were able to report the approximate date and location of vaccine receipt; participants were defined as unvaccinated if they reported that they had not received influenza vaccination in the current season; all other participants were considered to have unknown self-reported influenza vaccination status. Vaccination records were sought from each health system’s electronic medical records and MCIR for all participants regardless of self-reported vaccination status. Documentation of vaccine receipt for those who reported vaccination from a provider other than the enrolling health system was also requested from the specified provider.

Type of vaccine provider was based on self-reported vaccine location and categorized as health system of enrollment, out of system clinic, pharmacy, or work / community provider. Participants who reported that they were unvaccinated, but had a record of vaccination from the health system of enrollment were considered to have been vaccinated in that health system. Participants who reported that they were vaccinated in the health system of enrollment, but had no record of vaccination in that system’s records were considered to have been vaccinated at an unknown provider type.

### Statistical Analysis

Annual study populations were characterized by age group (<5, 5-19, 20-64, and ≥65 years), sex, race/ethnicity, health system of enrollment, and influenza vaccination determined by MCIR, health system, outside provider records, or self-report. The health system of enrollment is deidentified in the results and reported as Health System A or B.

The performance of MCIR in documenting influenza vaccination was assessed relative to vaccination documented by health system records, outside provider records, or self-report. Percent agreement, Cohen’s kappa, sensitivity, specificity, positive predictive value, and negative predictive values were calculated for each study year. The kappa statistic was interpreted as: 0-0.20 no agreement, 0.21-0.39, minimal; 0.40-0.59, weak; 0.60-0.79 moderate; 0.80-0.90 strong; and > 0.90 almost perfect [11]. These statistics were calculated overall, for adults ≥20 years, and for children <20 years for whom MCIR reporting is mandated.

Among subjects defined as vaccinated by combined provider records and self-report, the proportion documented by MCIR was compared by age group, sex, race/ethnicity, and vaccine provider type for each study year and trends over time assessed.

All statistical analysis was performed using SAS (version 9.4).

## RESULTS

In total, 11,503 participants were included in this analysis; annual enrollment varied from a low of 905 in 2013-2014 to a high of 1,945 in 2018-2019. Females consistently represented a slight majority of enrolled participants in each season. The distributions of age, race, and health system of enrollment varied from year to year (Table 1). Influenza vaccination coverage defined by any source (MCIR, health system or provider records, or self-report) increased over time from 45% in 2011-2012 to 66% in 2018-2019.

**Table 1.** Demographic characteristics of study participants by study year.

|  | 2011-2012<br>No. = 1425 | 2012-2013<br>No. = 1426 | 2013-2014<br>No. = 905 | 2014-2015<br>No. = 1587 | 2015-2016<br>No. = 1328 | 2016-2017<br>No. = 1365 | 2017-2018<br>No. = 1522 | 2018-2019<br>No. = 1945 |
| --- | --- | --- | --- | --- | --- | --- | --- | --- |
| Age |  |  |  |  |  |  |  |  |
| <5 years | 368 (25.8) | 309 (21.7) | 160 (17.7) | 311 (19.6) | 248 (18.7) | 269 (19.7) | 286 (18.8) | 293 (15.1) |
| 5-19 years | 427 (30) | 456 (32) | 180 (19.9) | 403 (25.4) | 404 (30.4) | 414 (30.3) | 402 (26.4) | 452 (23.2) |
| 20-64 years | 549 (38.5) | 569 (39.9) | 468 (51.7) | 697 (43.9) | 571 (43) | 569 (41.7) | 671 (44.1) | 933 (48) |
| ≥65 years | 81 (5.7) | 92 (6.5) | 97 (10.7) | 176 (11.1) | 105 (7.9) | 113 (8.3) | 163 (10.7) | 267 (13.7) |
| Sex |  |  |  |  |  |  |  |  |
| Female | 816 (57.3) | 799 (56) | 547 (60.4) | 927 (58.4) | 757 (57) | 788 (57.7) | 880 (57.8) | 1209 (62.2) |
| Male | 609 (42.7) | 627 (44) | 358 (39.6) | 660 (41.6) | 571 (43) | 577 (42.3) | 642 (42.2) | 736 (37.8) |
| Race / Ethnicity |  |  |  |  |  |  |  |  |
| Non-Hispanic White | 791 (55.5) | 801 (56.2) | 552 (61) | 1027 (64.7) | 894 (67.3) | 932 (68.3) | 987 (64.9) | 1272 (65.4) |
| Non-Hispanic Black | 350 (24.6) | 326 (22.9) | 190 (21) | 270 (17) | 192 (14.5) | 180 (13.2) | 205 (13.5) | 381 (19.6) |
| Asian | 68 (4.8) | 69 (4.8) | 44 (4.9) | 83 (5.2) | 57 (4.3) | 70 (5.1) | 121 (8) | 110 (5.7) |
| Hispanic | 86 (6) | 86 (6) | 54 (6) | 77 (4.9) | 58 (4.4) | 89 (6.5) | 84 (5.5) | 97 (5) |
| Other / Multiracial / Unknown | 130 (9.1) | 144 (10.1) | 65 (7.2) | 130 (8.2) | 127 (9.6) | 94 (6.9) | 125 (8.2) | 85 (4.4) |
| Health System of Enrollment |  |  |  |  |  |  |  |  |
| A | 619 (43.4) | 820 (57.5) | 506 (55.9) | 1045 (65.9) | 947 (71.3) | 958 (70.2) | 996 (65.4) | 1421 (73.1) |
| B | 806 (56.6) | 606 (42.5) | 399 (44.1) | 542 (34.2) | 381 (28.7) | 407 (29.8) | 526 (34.6) | 524 (26.9) |
| Influenza Vaccination<sup>1</sup> |  |  |  |  |  |  |  |  |
| Yes | 646 (45.3) | 746 (52.3) | 514 (56.8) | 980 (61.8) | 796 (59.9) | 775 (56.8) | 918 (60.3) | 1285 (66.1) |
| No | 779 (54.7) | 680 (47.7) | 391 (43.2) | 607 (38.3) | 532 (40.1) | 590 (43.2) | 604 (39.7) | 660 (33.9) |

<sup>1</sup> Defined by any source (MCIR, health system or provider records, or self-report)

Overall, the performance of MCIR in documenting influenza vaccination improved over time relative to combined provider records and self-report (Table 2). In the first four years of the study, there was moderate agreement between MCIR and combined provider records and self-report (Kappa range: 0.63-0.73). There was strong agreement in the last four years of the study (Kappa range: 0.83-0.86). In all years, the specificity and positive predictive value of MCIR records were very high reflecting that few vaccination records were identified by MCIR but not other sources. In contrast, the sensitivity and negative predictive value of MCIR records were lower reflecting that more than 10% of vaccinations identified by other sources were not captured by MCIR in each season. However, these measures did improve over time. The proportion of records not captured by MCIR decreased from a high of 33% in 2013-2014 to a low of 11% in 2018-2019. Over the same years, the sensitivity improved from 67% to 89% and the negative predictive value improved from 69% to 82%.

**Table 2.** Comparison of vaccination status defined by Michigan Care Improvement Registry (MCIR) relative to vaccination documented by health system records, outside provider records, or self-report.

|  | Vaccinated<sup>1</sup> | Unvaccinated<sup>1</sup> | Percent Agreement | Kappa | Sensitivity (95% CI) | Specificity (95% CI) | PPV (95% CI) | NPV (95% CI) |
| --- | --- | --- | --- | --- | --- | --- | --- | --- |
| 2011-2012 |  |  |  |  |  |  |  |  |
| MCIR vaccinated | 458 | 7 | 86.5 | 0.73 | 72.8 (69.2, 76.3) | 99.0 (97.9, 99.6) | 98.5 (96.9, 99.4) | 80.0 (77.2, 82.6) |
| MCIR unvaccinated | 171 | 684 |  |  |  |  |  |  |
| 2012-2013 |  |  |  |  |  |  |  |  |
| MCIR vaccinated | 537 | 2 | 85.3 | 0.71 | 72.7 (69.3, 75.9) | 99.7 (98.9, 100) | 99.6 (98.7, 100) | 76.2 (73.1, 79.0) |
| MCIR unvaccinated | 202 | 645 |  |  |  |  |  |  |
| 2013-2014 |  |  |  |  |  |  |  |  |
| MCIR vaccinated | 342 | 1 | 80.7 | 0.63 | 66.8 (62.5, 70.9) | 99.7 (98.5, 100) | 99.7 (98.4, 100) | 68.8 (64.7, 72.7) |
| MCIR unvaccinated | 170 | 375 |  |  |  |  |  |  |
| 2014-2015 |  |  |  |  |  |  |  |  |
| MCIR vaccinated | 762 | 0 | 86.3 | 0.73 | 78.2 (75.4, 80.7) | 100 (99.4, 100) | 100 (99.5, 100) | 73.0 (69.7, 76.0) |
| MCIR unvaccinated | 213 | 575 |  |  |  |  |  |  |
| 2015-2016 |  |  |  |  |  |  |  |  |
| MCIR vaccinated | 685 | 0 | 91.6 | 0.83 | 86.3 (83.7, 88.6) | 100 (99.3, 100) | 100 (99.5, 100) | 82.4 (79.2, 85.3) |
| MCIR unvaccinated | 109 | 511 |  |  |  |  |  |  |
| 2016-2017 |  |  |  |  |  |  |  |  |
| MCIR vaccinated | 675 | 1 | 93.1 | 0.86 | 88.2 (85.7, 90.4) | 99.8 (99.0, 100) | 99.9 (99.2, 100) | 86.0 (83.1, 88.6) |
| MCIR unvaccinated | 90 | 554 |  |  |  |  |  |  |
| 2017-2018 |  |  |  |  |  |  |  |  |
| MCIR vaccinated | 800 | 1 | 92.7 | 0.85 | 88.2 (85.9, 90.2) | 99.8 (99.0, 100) | 99.9 (99.3, 100) | 84.1 (81.1, 86.8) |
| MCIR unvaccinated | 107 | 565 |  |  |  |  |  |  |
| 2018-2019 |  |  |  |  |  |  |  |  |
| MCIR vaccinated | 1133 | 5 | 92.6 | 0.84 | 89.4 (87.5, 91.0) | 99.2 (98.1, 99.7) | 99.6 (99.0, 99.9) | 82.1 (79.2, 84.8) |
| MCIR unvaccinated | 135 | 620 |  |  |  |  |  |  |

Abbreviations: 95% CI, 95% confidence interval; PPV, positive predictive value; NPV, negative predictive value
<sup>1</sup> Defined by combined health system records, outside provider records, or self-report.

The performance of MCIR in documenting influenza vaccination defined by combined provider records and self-report varied by age and type of vaccine provider. For children 19 years and younger, for whom reporting is mandated, MCIR captured over 90% of vaccinations in most years (Figure 1). MCIR capture for adults was much lower in the first half of the study period, but approached levels observed for children by the end of the study period. MCIR capture for adults 20-64 years increased from 51% in 2010-2011 to 85% in 2018-2019, and capture for older adults increased from 33% in 2010-2011 to 93% in 2018-2019. The distribution of vaccine provider types remained relatively stable over time (Figure 2A), with vaccination in the enrolling health system representing the majority of vaccines received each year (range 75%-84%). MCIR consistently captured most (range 90%-97%) of the vaccinations given in Health System A each year (Figure 2B). Documentation in MCIR of vaccinations given in Health System B and from pharmacies increased over time with MCIR capturing over 90% of vaccination from these sources in 2018-2019. Relatively few vaccinations were received from sources outside the enrolling health systems, and as a result the percent of these vaccinations captured by MCIR was more variable from year to year. Nevertheless, some trends were observed. Capture of vaccinations received at work or other community sites increased over time, but peaking at only 61% in 2017-2018. In each year, there was moderate capture of vaccination from out of system clinics, and low capture of vaccinations from unknown locations with no consistent pattern over time. Capture of vaccinations by MCIR did not vary substantially by sex or race/ethnicity (Supplementary Figure 1).

**Figure 1.**
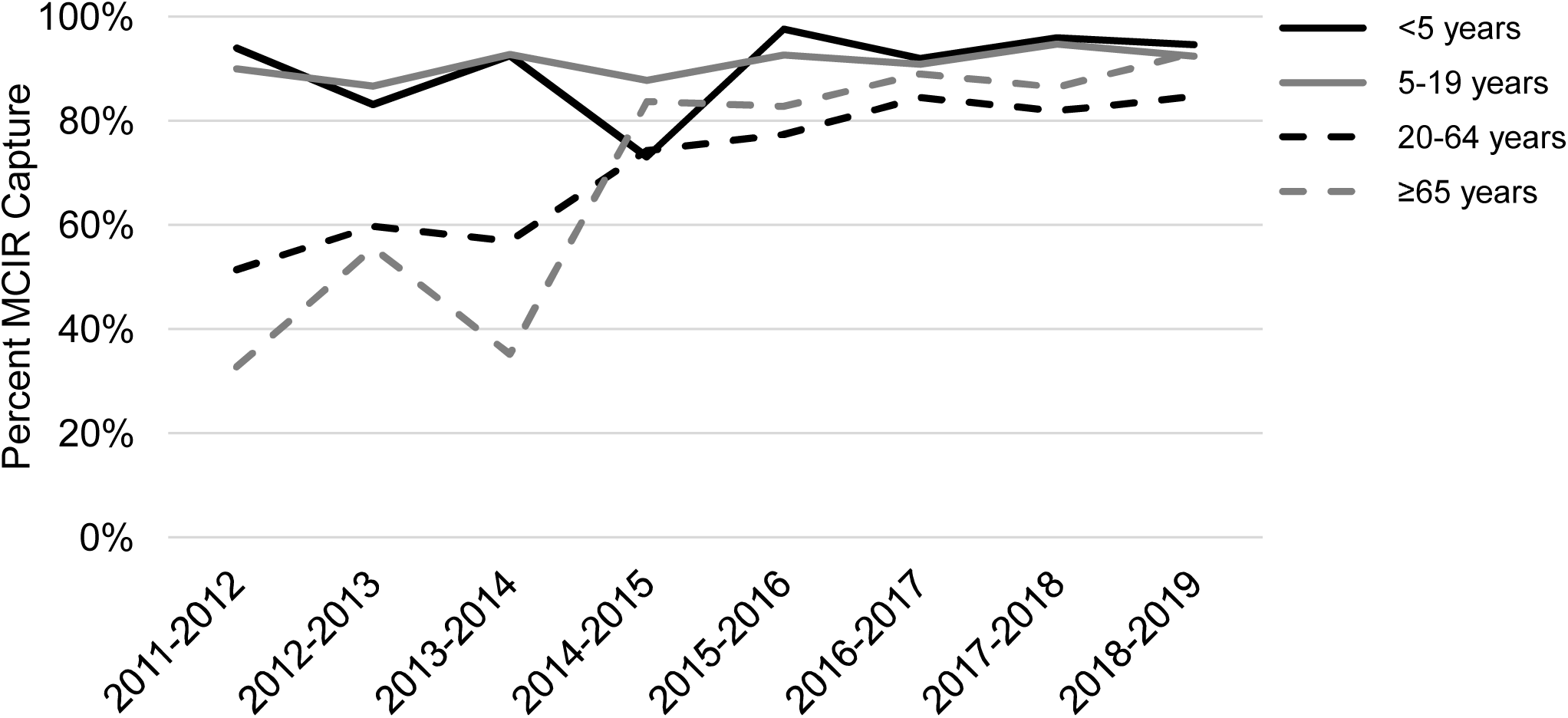
Percent of influenza vaccinations^1^ captured by the Michigan Care Improvement Registry (MCIR) by age: 2011-2019. ^1^Defined by combined health system records, outside provider records, or self-report.

**Figure 2.**
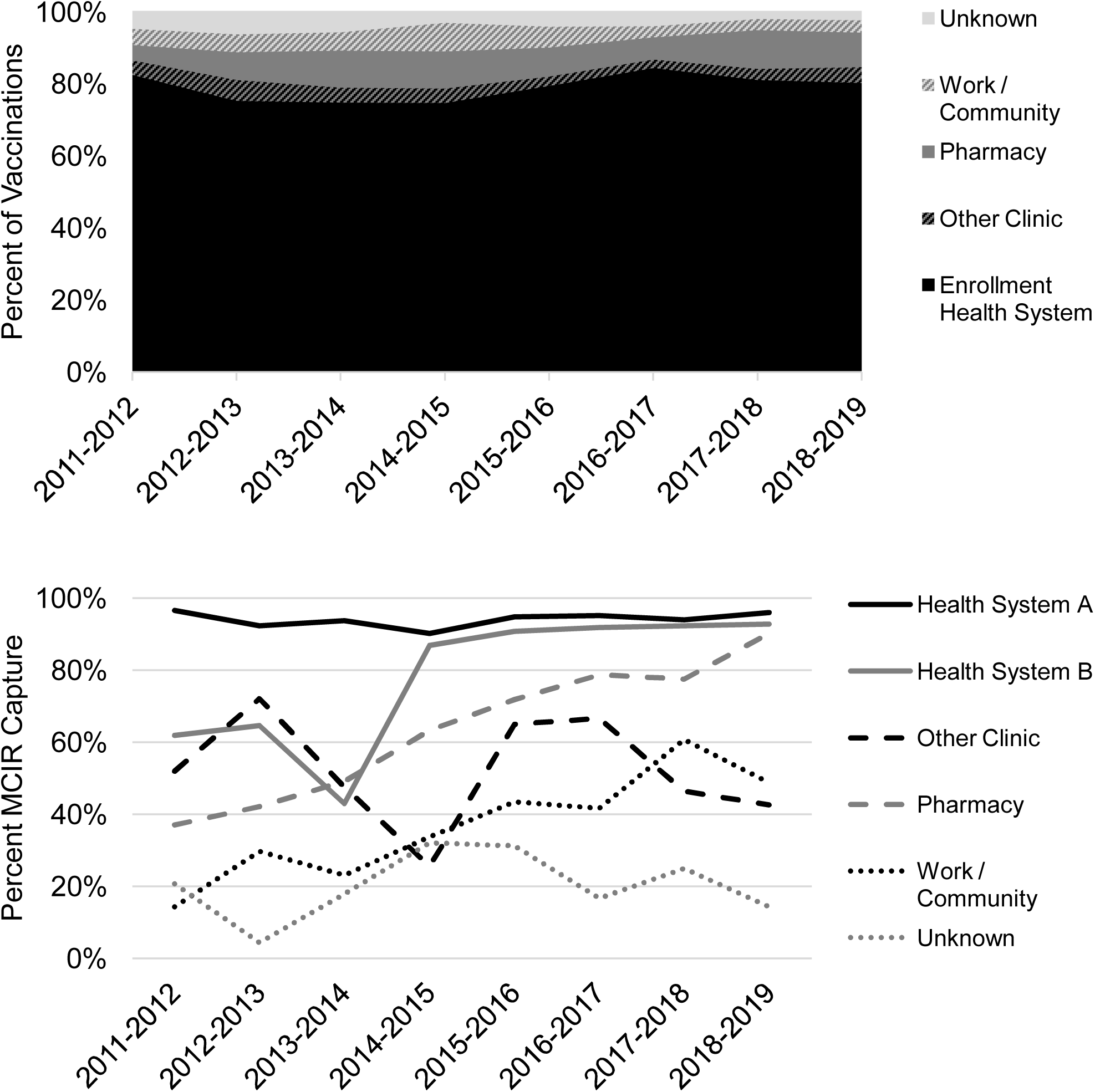
A) Proportion of influenza vaccinations given by provider type, and B) percent of influenza vaccinations captured by the Michigan Care Improvement Registry (MCIR) by provider type: 2011-2019.

## DISCUSSION

During our study period MCIR consistently captured of 90% of influenza vaccinations received by children <20 years of age for whom reporting is mandated. Overall, the performance of the registry improved over time. This improvement was largely driven by increased capture of adult vaccinations which approached that of children by 2018-2019 even though reporting was not required for adults during the entire study period. However, the sensitivity and negative predictive value of MCIR were <90% even in the most recent study year. This suggests that other sources including provider records and self-report ideally should continue to be used in combination with vaccine registries to reduce misclassification of vaccination status.

The mechanism of vaccine provider reporting seems to be an important factor in whether vaccines are captured in the registry. Health System B transitioned to a new electronic medical record software in the 2013-2014 season that facilitated automatic reporting to MCIR. After a decrease during this transition year, MCIR reporting from Health System B dramatically increased. This improved reporting from Health System B, along with continuous improvement in reporting from commercial pharmacies during the study period, drove the majority of improvement in reporting for adults. Reporting from work or community providers also consistently improved over the study period, but reporting remained lower than for the enrolling health systems and pharmacies. This likely reflects heterogeneity in the providers of vaccines at these locations. For example, some commercial pharmacies provide onsite vaccination services to large employers and these records may be reported through the pharmacy’s usual mechanisms. Despite overall increases, reporting from clinics outside the enrolling health systems, work or community providers, and other providers remains relatively low. These providers typically accounted for around 10% of the vaccines received in our study population, but likely account for a higher percentage vaccines provided nationally. According to the Centers for Disease Control and Prevention, 42.3% of adults receive their influenza vaccine at a non-medical setting [12]. This highlights the ongoing need to seek records from providers who do not routinely report to registries.

We studied the performance of MCIR in capturing records of influenza vaccination for participants enrolled from ambulatory clinics in 2 health systems. The performance of MCIR among these patients may not be generalizable to individuals who receive care from other health systems or those who do not have a primary care provider. In addition, while all 50 states have registries based on the Immunization Information System[13], the performance of vaccine registries is likely to vary by local reporting, legislation, and other factors. We compared influenza vaccination records from MCIR to vaccination defined by combined provider records and self-report. While previous studies have demonstrated self-report to be reliable, some level of misclassification in our definition of vaccination status is likely. As a result, we might be underestimating the performance of MCIR.

Assessment of the relative effectiveness of the various currently available influenza vaccine products is critical to informing their use in different groups to maximize population-level benefits. In addition, the next generation of influenza vaccines, developed in accordance with the National Institute of Allergy and Infectious Diseases strategic plan, will need to be evaluated in real world situations once they are licensed[14]. The two currently authorized SARS-CoV-2 vaccines and those still in development will need continued study following licensure to evaluate long-term protection and safety[15–17]. Vaccination registries that accurately document the exact product that is received by an individual are critical to these evaluations. The State of Michigan’s vaccine registry, MCIR, is a valuable resource for this information. Along with other registries nationally, it will useful in providing necessary information on vaccines deployed not only for children but for broad age groups.

## Data Availability

Data are available by request to the authors.

## Abbreviations

CDC: Centers for Disease Control and Prevention
CI: confidence interval
MCIR: Michigan Care Improvement Registry
US: United States
VE: vaccine effectiveness

## Acknowledgements

This work was supported by the Centers for Disease Control and Prevention (grant number U01 IP001034).

## Potential Conflicts of interest

E.T.M has received grant support from Merck and Pfizer for work unrelated to this report. A.S.M. has received consultancy fees from Sanofi and Seqirus for work unrelated to this report. All authors reported no potential conflicts.

**Supplemental Figure 1.**
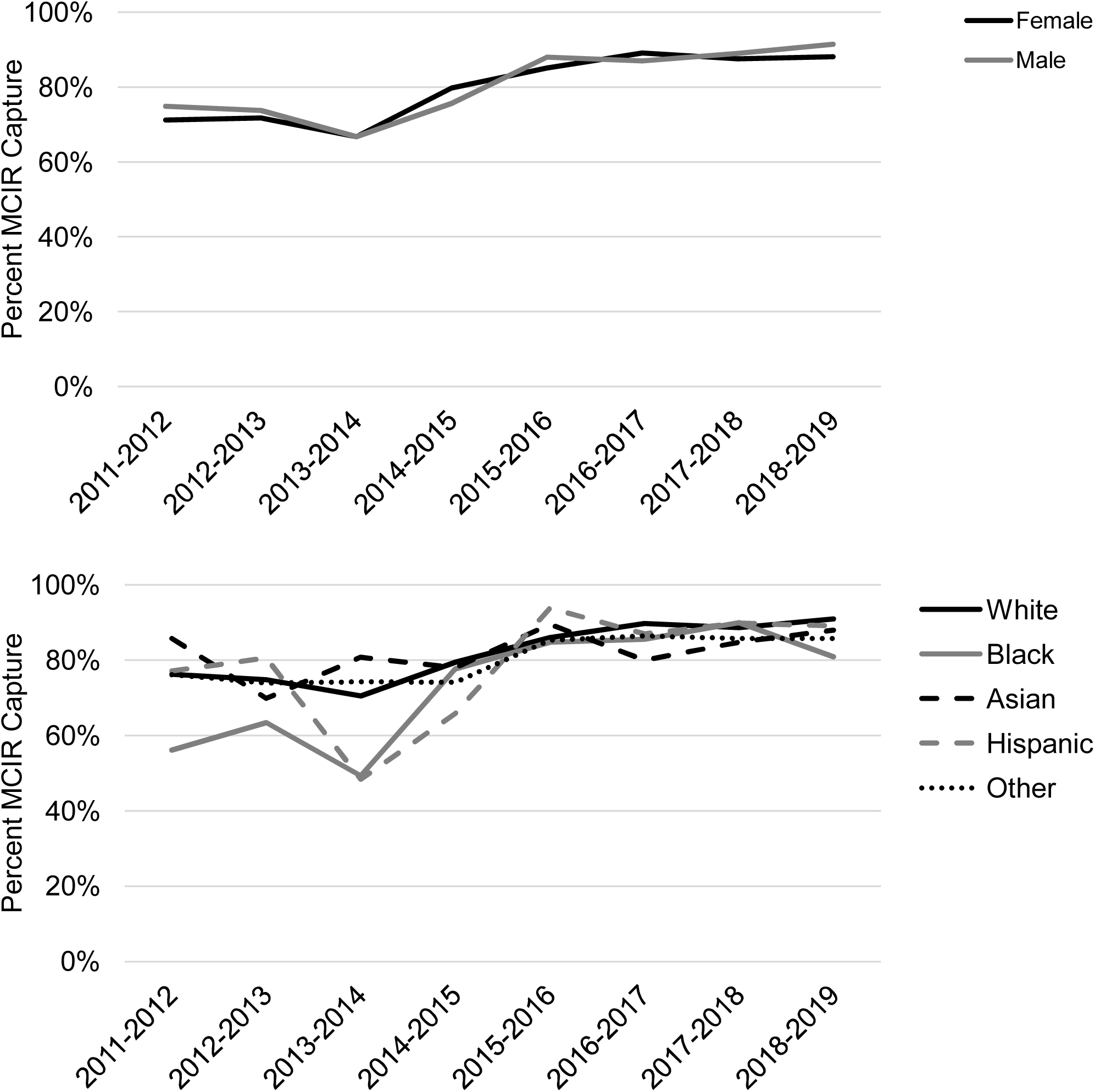
A) Percent of influenza vaccinations^1^ captured by the Michigan Care Improvement Registry by sex, and B) by race / ethnicity: 2011-2019. ^1^ Defined by combined health system records, outside provider records, or self-report.

